# Burden and risk factors for Achilles tendon rupture in the military population from 2006 to 2015: A retrospective cohort study

**DOI:** 10.1101/2023.10.02.23296425

**Authors:** John J. Fraser, Jennifer A. Zellers, Christopher K Sullivan, Cory F. Janney

## Abstract

**Background:** Achilles tendon rupture (ATR) is a serious musculoskeletal injury that results in substantial functional decline, especially in highly physically demanding occupations such as service in the military

**Purpose:** The objective of this study was to evaluate the burden and associated factors of ATR in US military service members.

**Study Design:** Descriptive Epidemiology Study

**Methods:** The Defense Medical Epidemiology Database was used to identify all diagnosed ATR in military personnel from 2006 to 2015. Prevalence of ATR was calculated and compared by year, service branch, and military rank. Unadjusted and adjusted assessments of injury risk were calculated.

**Results:** Officers incurred 15 978 episodes at a prevalence of 7.43/1000 (male: 8.11/1000; female: 3.89/1000). Among enlisted personnel, there were 59 242 episodes of ATR that occurred at a prevalence of 6.23 episodes per 1000 (male enlisted: 6.49/1000; female enlisted: 4.48/1000). Apart from enlisted aviation specialists (where there was no significant difference in risk between men and women), both female officers and enlisted service members had significantly lower risk of ATR compared with their male counterparts in all occupations (prevalence ratio [PR]: 0.26-0.73). Aviation and service officers demonstrated significantly lower risk of ATR episodes (PR: 0.87-0.91) and administration, operations, intelligence, and logistic officers demonstrated increased risk (PR: 1.16-1.31) compared with ground and naval gunfire officers. Among enlisted specialties, all but mechanized/armor and combat engineers had significantly higher risk of ATR risk compared with infantry (PR: 1.14-2.13), with the highest risk observed in the administration, intelligence, and communication fields.

**Conclusions:** ATR was ubiquitous in the US military, with multiple risk factors identified, including male sex, older age, rank, military occupation, and service branch.

**Clinical Relevance:** Based on the burden of ATR in the US military observed in this study, these findings highlight both the need for prophylactic interventions and identification of the populations who can most greatly benefit from preventive screening and care.

**What is known about the subject:** In an earlier study of the ATR burden in US military members, 1441 ATR cases were identified between 1998 and 2001, occurring at a rate of 30.9 per 100 000 person-years. In a more recent study of care episodes for Achilles tendinopathy in the military, the prevalence was 17.65 per 1000 in officers and 12.22 per 1000 in enlisted members, with male sex, older age, senior rank, military occupation, and service branch found to be salient associated factors. It is highly plausible that occupation may also be a salient factor for ATR, given that tendinopathy may be a prodromal sign of future tendon failure.

**What this study adds to existing knowledge:** Due to the changes in operational requirements, training, engagement in overseas contingency operations, and force composition that have occurred over the past 22 years, this provides an updated assessment of burden and the associated risk factors of ATR.

## INTRODUCTION

The Achilles tendon, also known as the calcaneal tendon, has an important role in propulsion, force attenuation, and energy return during walking, running, and jumping during function. Achilles tendon rupture (ATR) is a serious musculoskeletal injury that results in substantial functional decline, especially in highly physically demanding occupations such as service in the military.^1^ In the general public, ATR affects an estimated 32 906 individuals in the US per year, at an overall rate of 2.1 per 100 000 person-years.^2^ Some of the risk factors associated with ATR include previous musculoskeletal injury, chronic Achilles tendon inflammation and tendinopathy, regular participation in athletic activity, spring season, genetic factors, chronic systemic conditions (diabetes, hyperparathyroidism, and renal failure), and medication use (steroids, quinolones, and oral bisphosphonate).^3^ ATR management includes both conservative and surgical approaches, with return to sport occurring in 80% of affected athletes^4^ and return to duty in 90-100% of affected military members.^1, 5^ Among military members, rehabilitation takes an average of 8.2 months in nonoperative cases^1^ and 5.4 to 6.7 months following surgery^1, 5^ before they are cleared for return to full, unrestricted duty.

In an earlier study of the ATR burden in US military members, 1441 ATR cases were identified between 1998 and 2001, occurring at a rate of 30.9 per 100 000 person-years.^6^ The 15-fold higher burden in the military may in part be due to the unique exposures incurred during military service. In a more recent study of care episodes for Achilles tendinopathy in the military, the prevalence was 17.65 per 1000 in officers and 12.22 per 1000 in enlisted members, with male sex, older age, senior rank, military occupation, and service branch found to be salient associated factors.^7^ It is highly plausible that occupation may also be a salient factor for ATR, given that tendinopathy may be a prodromal sign of future tendon failure. Due to the changes in operational requirements, training, engagement in overseas contingency operations, and force composition that have occurred over the past 22 years, an updated assessment of burden and the associated risk factors of ATR is warranted. Therefore, the purpose of this study was to determine the burden of ATR in the military population with consideration of military occupation and identify risk factors that may contribute to this condition.

## METHODS

A population-based epidemiological retrospective cohort study of all service members in the US Armed Forces was performed, assessing sex, military occupation, rank, branch, and year on ATR prevalence from 2006 to 2015. The Defense Medical Epidemiology Database (DMED; Defense Health Agency, Falls Church, VA, https://bit.ly/DHADMED5) was utilized to identify relevant healthcare encounters. This database provides aggregated data for International Classification of Diseases, Ninth Revision, Clinical Modification (ICD-9-CM) codes and de-identified patient characteristics, including sex, categories of military occupations, and branch of service for all active duty and reserve military service members. The database is HIPAA compliant, does not include any personally identifiable or personal health information, and has been used previously for the epidemiological study of ankle-foot injury in the military.^8, 9^ This study was approved as non–human-subjects research by the Institutional Review Board 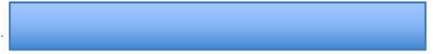 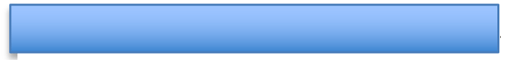. The Strengthening the Reporting of Observational Studies in Epidemiology (STROBE) guidelines^10^ were used to guide reporting.

The database was queried for the number of distinct care episodes for a primary diagnosis of ATR (ICD-9-CM codes 727.67, nontraumatic rupture of the Achilles tendon, and 845.09, other sprains and strains of ankle) from 2006 to 2015. Prevalence of ATR care episodes was calculated for male and female military members, enlisted and officers, in each service branch (Army, Navy, Marine Corps, and Air Force) and occupational category.^8, 9^ Prevalence ratio (PR) point estimates and 95% confidence intervals (CIs), risk difference point estimates, attributable risk, number needed to harm, and chi-square statistics were calculated in the assessment of sex and occupation category using Microsoft Excel for Mac 2016 (Microsoft Corp., Redmond, WA) and a custom epidemiological spreadsheet.^11^ In the unadjusted assessment of sex and occupation on ATR risk, male service members, enlisted infantry, and ground/naval gunfire officer groups were used as the reference categories.

Due to overdispersion of the data, a negative binomial model was employed over a Poisson model for the adjusted assessment of female sex (male reference), branch (US Army reference), officer rank (enlisted reference), and year (2006 reference) on the risk of ATR.^12^ Since there were excess zeros noted in the data, with many demographic categories not having any reported outcomes of interest (especially among the smaller subpopulations), assessment of convergence between the unadjusted and adjusted (hurdle) negative binomial regression models were performed.^7^ In the hurdle negative binomial regression model, a logistic link is employed to remove demographic categories that contributed no episodes of ATR from the final count model. By doing so, excess zeros that contribute to skewed point estimates, standard errors, and overdispersion are parsed in the truncated regression model.^12^ Results of hurdle negative binomial regression are reported using calculations of the predictors regressed on count data (count model assessing the rate of the outcome) as well as a linked logistic regression (zero model that assessed the probability within demographic categories of not having the outcome).

Both unadjusted and adjusted models are reported in the supplemental material. The regression analyses were performed using the PSCL (version 1.5) and MASS (version 7.3-58.1) packages on R (version 3.5.1, The R Foundation for Statistical Computing, Vienna, Austria). The level of significance was *p* < 0.05 for all analyses. PR point estimates were considered statistically significant if CIs did not cross the 1.00 threshold. Convergence of *p* values, effect sizes, and 95% CIs were considered when evaluating significant findings.

## RESULTS

**Supplemental tables 1 and 2** detail the counts and prevalence of ATR in officer and enlisted personnel. In the study epoch, officers incurred 15 978 episodes of ATR at a prevalence of 7.43 per 1000 (male officers: 8.11 per 1000; female officers: 3.89 per 1000). Among enlisted personnel, 59 242 ATR episodes occurred at a prevalence of 6.23 episodes per 1000 (male enlisted: 6.49 per 1000; female enlisted: 4.48 per 1000). **Table 1** details the univariate factor of sex within occupational specialty on ATR episodes. Apart from enlisted aviation specialists (with no significant difference in risk was between men and women), both female officers and enlisted service members had significantly lower risk of ATR compared with their male counterparts in all occupations (PR: 0.26-0.73).

**TABLE 1.**
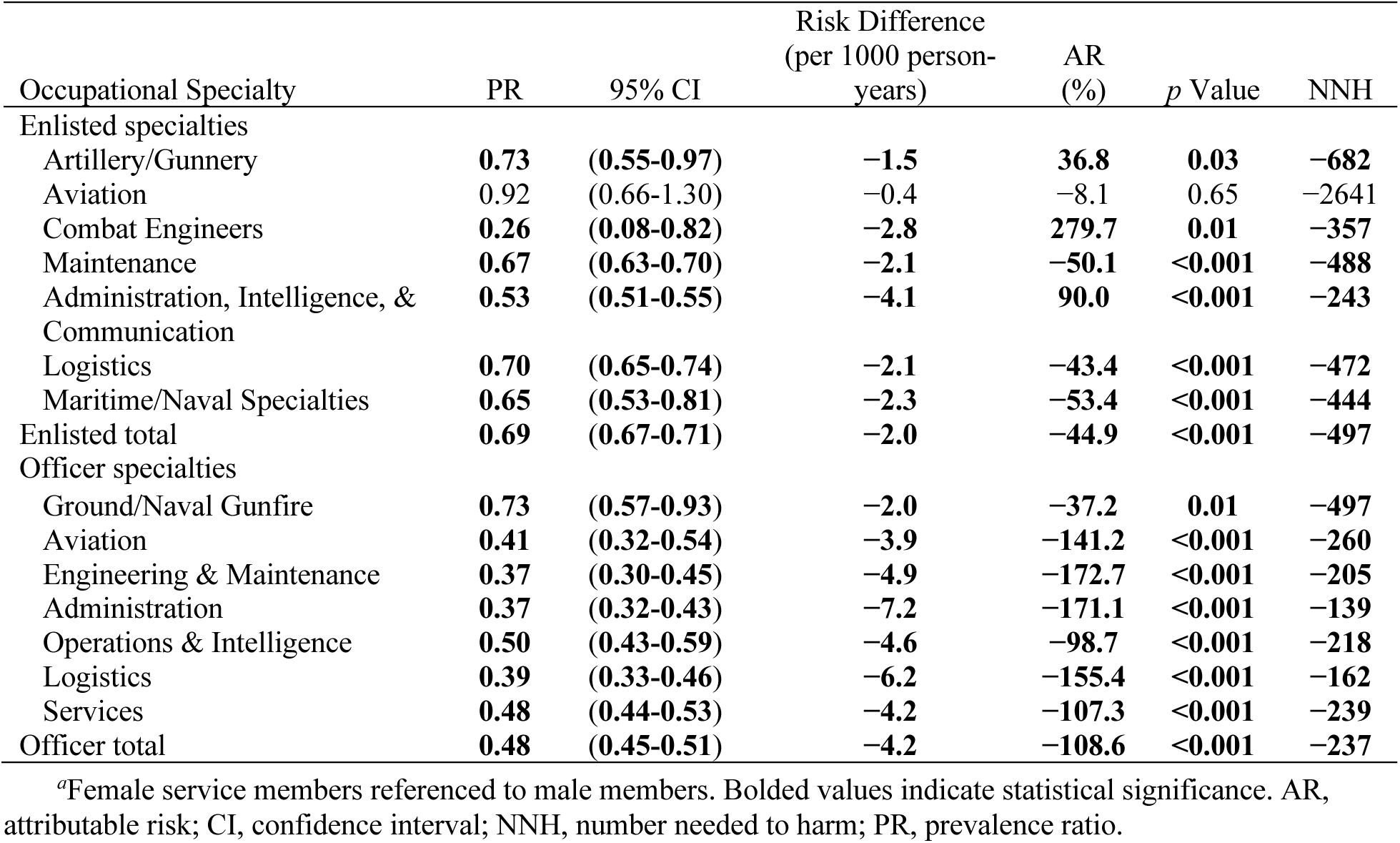
Assessment of Risk of Achilles Tendon Rupture by Sex Within Occupation in Members of the US Armed Forces, 2006-2015*^a^*.

In the assessment of occupational exposure, aviation and service officers demonstrated significantly lower risk of ATR episodes (PR: 0.87-0.91) and administration, operations, intelligence, and logistic officers demonstrated increased risk (PR: 1.16-1.31) compared with ground and naval gunfire officers (**Table 2**). There was no significant difference in PR between engineering and maintenance officers and the reference group. Among enlisted specialties, all but mechanized/armor and combat engineers had significantly higher ATR risk compared with infantry (PR: 1.14-2.13), with the highest risk observed in the administration, intelligence, and communication fields.

**TABLE 2.** Assessment of Risk of Achilles Tendon Rupture by Occupation in the US Armed Forces, 2006-2015*^a^*.

| Occupational Specialty | PR | 95% CI | Risk Difference<br>(per 1000 person-years) | AR<br>(%) | *p* value | NNH |
| --- | --- | --- | --- | --- | --- | --- |
| Enlisted specialties<sup>b</sup> |  |  |  |  |  |  |
| Special Operations Forces | **1.14** | **(1.01-1.28)** | **0.5** | **12.0** | **0.03** | **2024** |
| Mechanized/Armor | 1.03 | (0.93-1.14) | 0.1 | 3.1 | 0.55 | 8635 |
| Artillery/Gunnery | **1.48** | **(1.40-1.57)** | **1.8** | **32.5** | **<0.001** | **571** |
| Aviation | **1.38** | **(1.26-1.51)** | **1.4** | **27.6** | **<0.001** | **724** |
| Combat Engineers | 1.03 | (0.94-1.13) | 0.1 | 3.1 | 0.48 | 8580 |
| Maintenance | **1.64** | **(1.59-1.70)** | **2.3** | **39.1** | **<0.001** | **428** |
| Administration, Intelligence, &<br>Communication | **2.13** | **(2.05-2.20)** | **4.1** | **53.0** | **<0.001** | **244** |
| Logistics | **1.83** | **(1.76-1.90)** | **3.0** | **45.4** | **<0.001** | **331** |
| Maritime/Naval Specialties | **1.67** | **(1.54-1.80)** | **2.4** | **40.0** | **<0.001** | **412** |
| Officer specialties<sup>c</sup> |  |  |  |  |  |  |
| Aviation | **0.87** | **(0.82-0.92)** | **-1.0** | **-15.0** | **<0.001** | **-1041** |
| Engineering & Maintenance | 0.97 | (0.91-1.03) | -0.2 | -3.2 | 0.27 | -4336 |
| Administration | **1.31** | **(1.23-1.39)** | **2.2** | **23.4** | **<0.001** | **446** |
| Operations & Intelligence | **1.16** | **(1.09-1.23)** | **1.2** | **13.7** | **<0.001** | **860** |
| Logistics | **1.22** | **(1.15-1.30)** | **1.6** | **18.1** | **<0.001** | **617** |
| Services | **0.91** | **(0.87-0.96)** | **-0.6** | **-9.5** | **<0.001** | **-1571** |

<sup>a</sup>Bolded values indicate statistical significance. AR, attributable risk; CI, confidence interval; NNH, number needed to harm; PR, prevalence ratio.
<sup>b</sup>Referenced to enlisted infantry.
<sup>c</sup>Referenced to ground and naval gunfire officers.

**Table 3** details the results of the hurdle negative binomial regression model (adjusted for zero inflation) assessing sex, age range, rank, branch, and year on ATR risk. When controlling for other factors, female sex and senior officers demonstrated significantly lower PR of ATR than the male and junior enlisted reference groups, respectively. Older age (>30 years) and service in the US Marine Corps and Air Force were salient associated factors for increased risk of ATR (PR: 1.17-2.50). **Supplemental table 3** compares the negative binomial models with and without adjustment for zero inflation. While the results converged between the models for most of the variables assessed, the hurdle model outperformed the standard negative binomial model.

**TABLE 3.**
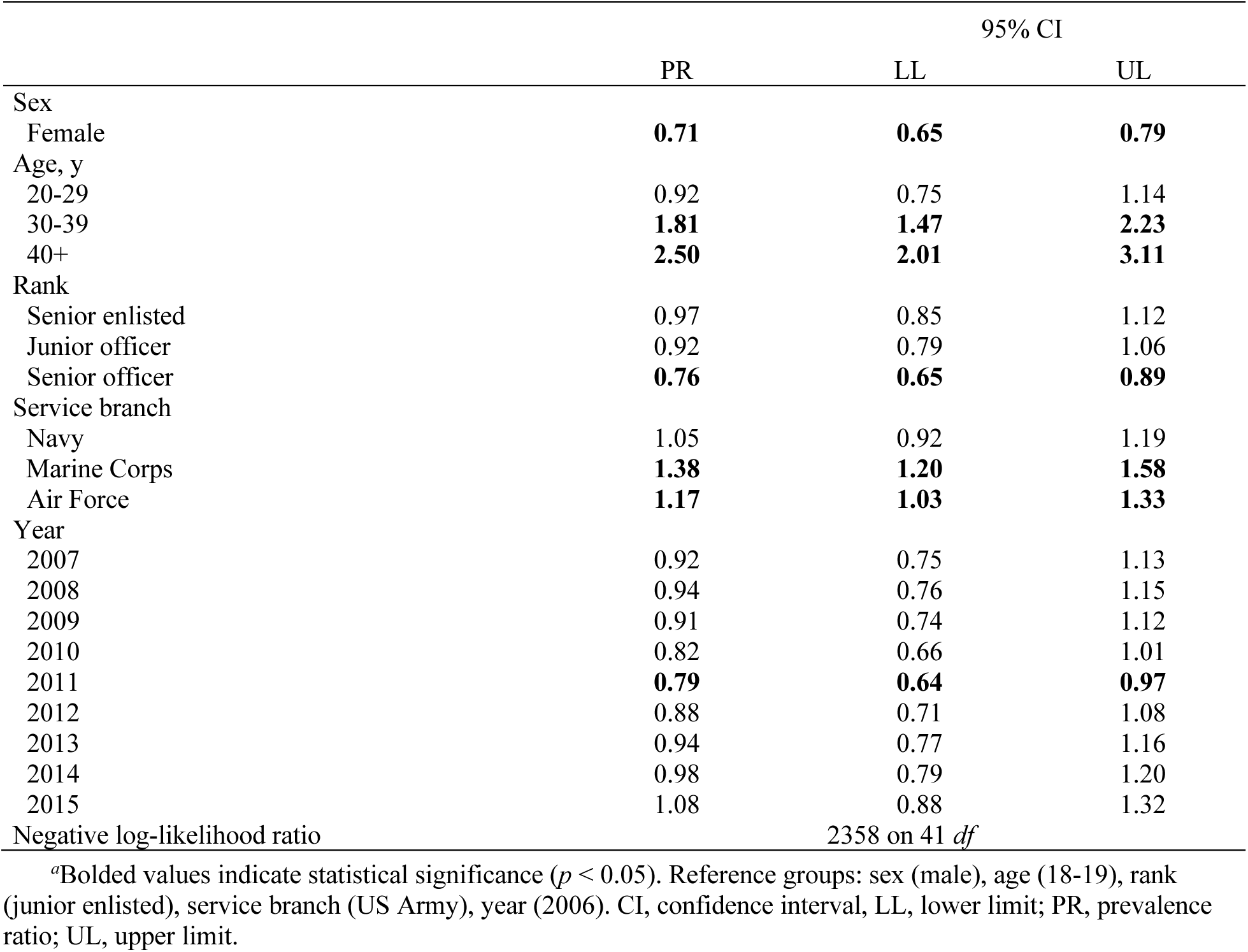
Negative Binomial Regression (Adjusted for Zero Inflation) Assessing Sex, Age, Rank, Service Branch, and Year on Prevalence of Achilles Tendon Rupture in the US Armed Forces, 2006-2015*^a^*.

## DISCUSSION

This study found that ATRs were common in the US military and identified multiple salient associated risk factors, including male sex, older age, enlisted or junior officer rank, military occupation, and service in the US Marine Corps or Air Force. Among military occupations, administration, operations, intelligence, and logistics specialties had the greatest risk for ATR in both officer and enlisted personnel. Integrating our understanding from the literature on civilian populations with the present study presents unique opportunities to interrogate specific aspects of mechanisms underlying ATR risk, including the contribution of exposure to activities with high Achilles tendon loads, sex, and age.

We found the burden of ATR more closely aligns with those previously described in the civilian population,^2^ contrasted with the high rates previously reported in the US military.^6^ The annual burden was consistent throughout the study epoch, despite the changes in exposure and tempo of military operations. This finding diverges from studies in the civilian population describing a progressive increased burden of ATR beginning in the 1990s and continuing to the present.^13^ It may be that an increasingly sedentary civilian population engaged in irregular bouts of activity (“weekend warriors”) has led to increased burden of ATR over time compared with the more consistently physically active military population.

Findings from this study underscore the importance of activity exposure on ATR risk. Interestingly, this study supports the framework that both overloading and underloading with bouts of high loads can be detrimental to Achilles tendon health. It may seem contradictory that service in the US Marine Corps (routinely encountering high Achilles tendon loading activities) and Air Force (routinely involved in low Achilles tendon loading activities) both had increased risk of Achilles tendon rupture compared with the US Army. We posit that these two branches represent different occupational risk exposures. Individuals in the US Marine Corps are routinely engaged in high-load running and jumping tasks and may represent an “overload” mechanism contribution to ATR risk. Alternatively, individuals in the Air Force are more routinely involved in technical occupations with longer bouts of sitting interspersed with less-frequent bouts of high Achilles loading activities. This group may represent a more traditional, “underload with intermittent high loading” mechanism contributing to ATR risk. Similarly, there is a lower risk of ATR in senior officers (when accounting for age), supporting the idea that individuals with high administrative loads, generally associated with longer bouts of sedentary activities without intermittent high loading bouts, have less occupational exposure to the high eccentric loads, leading to ATR.

A higher burden of ATR in men than in women has been well established in the civilian population, with injuries occurring 3-7 times more frequently in men.^14^ The mechanisms underlying this sexual dimorphism is a point of ongoing study. Studies of preclinical models of the Achilles tendon have found regulation of female hormones to be an important contributor to tendon mechanical behavior in the context of maintaining homeostasis and healing.^15, 16^ Furthermore, women have been found to have greater passive dorsiflexion motion compared with men, a function of triceps surae extensibility.^17^ Recent studies in human tendon have identified the presence of estrogen receptors in tendon tissue and estrogen effects on tendon biomechanical function, supporting the notion of sex hormone-mediated signaling in tendon.^18–20^ Studies investigating sex-mediated differences in patient outcomes after ATR have found conflicting results, though the propensity for male patients to incur this injury more than female patients is highly consistent across studies.^21–23^

The influence of sex on tendon homeostasis and healing in humans, though, is potentially confounded by a host of factors outside of direct hormonal influence. For example, it is possible that behavioral differences between sexes relating to engaging in recreational sport activity^24^ could partially account for differences in ATR burden, given that rupture tends to be associated with forceful eccentric contractions encountered during team sports like basketball and football. The present study informs this conversation by demonstrating that burden of ATR is higher in men than women when occupational exposure is comparable (for every occupational specialty included in this study, except for enlisted aviation). However, the relative risk encountered by women compared with men was lowest in specialties in which exposures are generally considered to have lower physical demands and are more sedentary (e.g., officer specialties, particularly administrative and logistics roles). The findings of this study add support to the notion that there is a mechanism by which sex influences ATR risk, and this risk is also influenced by repeated exposure to activities in which the Achilles tendon is likely to experience higher loads.

Service members above the age of 30 were found to be at increased risk of ATR. This finding aligns well with prior studies of ATR in civilian and elite athlete populations, where the highest incidence occurs in individuals aged 30-50 years.^13, 14, 25, 26^ Individuals in middle and older age groups have also consistently been found to have less optimal functional outcomes compared with younger individuals,^21, 23^ though individuals over the age of 65 have been reported to have greater improvements in self-reported postinjury outcomes compared with younger age groups.^22^ Identifying mechanisms underlying the isolated factor of age on ATR risk is challenging because it is often difficult to disentangle behavioral changes (increased sedentary behavior) and tendon maturation (age-related mechanical and physiological changes).^27^ That said, it does seem that aging can have an effect on tendon cellular function and matrix composition, though regular activity mitigates many of these changes.^27^ Similarly, aging has inconsistently been associated with reduced muscle cross-sectional area and altered Achilles tendon mechanical behavior, but these characteristics also seem to be moderated by physical activity level.^27, 28^ Reductions in muscle cross-sectional area and Achilles tendon extensibility, findings typically associated with aging, were not observed when participants were matched for height, weight, and activity level.^29^ The findings from the present study further support that the risk of ATR does not likely reflect aging alone but rather the combined effects of both age and exposure.

A substantial strength of this study was that using DMED allowed for assessment of population-level differences in ATR across several strata, including sex, occupation, military rank, branch of service, and year. This study also had some limitations. Using diagnostic codes has inherent weaknesses, especially in military populations where many determinants can influence care seeking.^30^ The data used in this study only represent individuals who sought treatment for their injuries and do not include individuals who self-managed their condition, though ATR presents with substantial functional deficits that would be difficult to self-manage, mitigating concerns regarding care-seeking behavior. This study also included each occurrence of ATR as a separate care episode. Individuals with a history of ATR have been found to have increased burden of rupture on the contralateral limb compared with the general population,^31^ so it is possible that a single person may have contributed more than one episode of care in the present study. Lastly, although personnel were categorized by their military occupation, a detailed task analysis to determine the specific duties and exposures that may be associated with these injuries was not possible.

## CONCLUSION

ATR was ubiquitous in the US military, with multiple risk factors identified, including male sex, older age, rank, military occupation, and service branch. Based on the burden of ATR in the US military observed in this study, these findings highlight both the need for prophylactic interventions and identification of the populations who can most greatly benefit from preventive screening and care.

## Data Availability

All data produced in the present work are contained in the manuscript

## REFERENCES

1. Renninger CH, Kuhn K, Fellars T, Youngblood S, Bellamy J. Operative and Nonoperative Management of Achilles Tendon Ruptures in Active Duty Military Population. Foot Ankle Int. 2016;37(3):269–273. doi:10.1177/1071100715615322

2. Lemme NJ, Li NY, DeFroda SF, Kleiner J, Owens BD. Epidemiology of Achilles Tendon Ruptures in the United States: Athletic and Nonathletic Injuries From 2012 to 2016. Orthop J Sports Med. 2018;6(11):2325967118808238. doi:10.1177/2325967118808238

3. Xergia SA, Tsarbou C, Liveris NI, Hadjithoma Μ, Tzanetakou IP. Risk factors for Achilles tendon rupture: an updated systematic review. Phys Sportsmed. Published online June 10, 2022:1–11. doi:10.1080/00913847.2022.2085505

4. Zellers JA, Carmont MR, Silbernagel KG. Return to Play Post Achilles Tendon Rupture: A Systematic Review and Meta-Analysis of Rate and Measures of Return to Play. Br J Sports Med. 2016;50(21):1325–1332. doi:10.1136/bjsports-2016-096106

5. Tropf J, Piscoya AS, Lundy A, Nelson S, Eckel TT. No Difference in Return to Duty Between Minimally Invasive, Percutaneous Achilles Tendon Repair and Nonoperative Functional Rehabilitation in Active Duty Military Members. Foot Ankle Orthop. 2022;7(2):2473011421S00544. doi:10.1177/2473011421S00544

6. Lum G. Spontaneous Ruptures of the Achilles Tendon, US Armed Forces, 1998-2001. Med Surveill Mon Rep. 2002;8.

7. Sullivan CK, Janney CF, Fraser JJ. Burden and risk factors for Achilles tendinopathy in the military population from 2006 to 2015. A retrospective cohort study. J Athl Train. 2023;(In Press).

8. MacGregor AJ, Fogleman SA, Dougherty AL, Ryans CP, Janney CF, Fraser JJ. Sex Differences in the Incidence and Risk of Ankle–Foot Complex Stress Fractures Among U.S. Military Personnel. J Womens Health. 2022;31(4):586–592. doi:10.1089/jwh.2021.0292

9. Fraser JJ, MacGregor AJ, Ryans CP, Dreyer MA, Gibboney MD, Rhon DI. Sex and occupation are salient factors associated with lateral ankle sprain risk in military tactical athletes. J Sci Med Sport. 2021;24(7):677–682. doi:10.1016/j.jsams.2021.02.016

10. von Elm E, Altman DG, Egger M, Pocock SJ, Gøtzsche PC, Vandenbroucke JP. The Strengthening the Reporting of Observational Studies in Epidemiology (STROBE) Statement: Guidelines for reporting observational studies. Int J Surg. 2014;12(12):1495–1499. doi:10.1016/j.ijsu.2014.07.013

11. LaMorte WW. Epidemiology/Biostatistics Tools [Excel workbook]. Published online 2006. Accessed August 3, 2019. http://bit.ly/Lamort3

12. Feng CX. A comparison of zero-inflated and hurdle models for modeling zero-inflated count data. J Stat Distrib Appl. 2021;8(1):8. doi:10.1186/s40488-021-00121-4

13. Sheth U, Wasserstein D, Jenkinson R, Moineddin R, Kreder H, Jaglal SB. The epidemiology and trends in management of acute Achilles tendon ruptures in Ontario, Canada: a population-based study of 27 607 patients. Bone Jt J. 2017;99-B(1):78–86. doi:10.1302/0301-620X.99B1.BJJ-2016-0434.R1

14. Lantto I, Heikkinen J, Flinkkilä T, Ohtonen P, Leppilahti J. Epidemiology of Achilles tendon ruptures: increasing incidence over a 33-year period. Scand J Med Sci Sports. 2015;25(1):e133–138. doi:10.1111/sms.12253

15. Fryhofer GW, Freedman BR, Hillin CD, et al. Postinjury biomechanics of Achilles tendon vary by sex and hormone status. J Appl Physiol. 2016;121(5):1106–1114. doi:10.1152/japplphysiol.00620.2016

16. Pardes AM, Freedman BR, Fryhofer GW, Salka NS, Bhatt PR, Soslowsky LJ. Males have Inferior Achilles Tendon Material Properties Compared to Females in a Rodent Model. Ann Biomed Eng. 2016;44(10):2901–2910. doi:10.1007/s10439-016-1635-1

17. Riemann BL, DeMont RG, Ryu K, Lephart SM. The Effects of Sex, Joint Angle, and the Gastrocnemius Muscle on Passive Ankle Joint Complex Stiffness. J Athl Train. 2001;36(4):369–375.

18. Bryant AL, Clark RA, Bartold S, et al. Effects of estrogen on the mechanical behavior of the human Achilles tendon in vivo. J Appl Physiol. 2008;105(4):1035–1043. doi:10.1152/japplphysiol.01281.2007

19. Leblanc DR, Schneider M, Angele P, Vollmer G, Docheva D. The effect of estrogen on tendon and ligament metabolism and function. J Steroid Biochem Mol Biol. 2017;172:106–116. doi:10.1016/j.jsbmb.2017.06.008

20. Astill BD, Katsma MS, Cauthon DJ, et al. Sex-based difference in Achilles peritendinous levels of matrix metalloproteinases and growth factors after acute resistance exercise. J Appl Physiol. 2017;122(2):361–367. doi:10.1152/japplphysiol.00878.2016

21. Carmont MR, Zellers JA, Brorsson A, Nilsson-Helander K, Karlsson J, Grävare Silbernagel K. Age and tightness of repair are predictors of heel-rise height after Achilles tendon rupture. Orthop J Sports Med. 2020;8(3):2325967120909556.

22. Cramer A, Jacobsen NC, Hansen MS, Sandholdt H, Hölmich P, Barfod KW. Outcome after acute Achilles tendon rupture is not negatively affected by female sex and age over 65 years. Knee Surg Sports Traumatol Arthrosc. 2020;28(12):3994–4002. doi:10.1007/s00167-020-06003-7

23. Olsson N, Petzold M, Brorsson A, Karlsson J, Eriksson BI, Grävare Silbernagel K. Predictors of clinical outcome after acute Achilles tendon ruptures. Am J Sports Med. 2014;42(6):1448–1455.

24. Rosenfeld CS. Sex-dependent differences in voluntary physical activity. J Neurosci Res. 2017;95(1-2):279–290. doi:10.1002/jnr.23896

25. Parekh SG, Wray WH, Brimmo O, Sennett BJ, Wapner KL. Epidemiology and Outcomes of Achilles Tendon Ruptures in the National Football League. Foot Ankle Spec. 2009;2(6):283–286. doi:10.1177/1938640009351138

26. Houshian S, Tscherning T, Riegels-Nielsen P. The epidemiology of achilles tendon rupture in a Danish county. Injury. 1998;29(9):651–654. doi:10.1016/S0020-1383(98)00147-8

27. Magnusson DM, Eisenhart M, Gorman I, Kennedy VK, E. Davenport T. Adopting Population Health Frameworks in Physical Therapist Practice, Research, and Education: The Urgency of Now. Phys Ther. 2019;99(8):1039–1047. doi:10.1093/ptj/pzz048

28. Lazarus NR, Harridge SDR. Declining performance of master athletes: silhouettes of the trajectory of healthy human ageing? J Physiol. 2017;595(9):2941–2948. doi:10.1113/JP272443

29. Couppé C, Svensson RB, Grosset JF, et al. Life-long endurance running is associated with reduced glycation and mechanical stress in connective tissue. Age Dordr Neth. 2014;36(4):9665. doi:10.1007/s11357-014-9665-9

30. Fraser JJ, Schmied E, Rosenthal MD, Davenport TE. Physical therapy as a force multiplier: population health perspectives to address short-term readiness and long-term health of military service members. Cardiopulm Phys Ther J. 2020;31(1):22–28. doi:10.1097/CPT.0000000000000129

31. Arøen A, Helgø D, Granlund OG, Bahr R. Contralateral tendon rupture risk is increased in individuals with a previous Achilles tendon rupture. Scand J Med Sci Sports. 2004;14(1):30–33. doi:10.1111/j.1600-0838.2004.00344.x

