## Supplemental table for "Burden and risk factors for Achilles tendon rupture in the military population from 2006 to 2015: A retrospective cohort study"

Supplemental Table 1. Count and prevalence of Achilles tendon ruptures among officers in the US Armed Forces, 2006-2015

| Officer Specialty | Ground and Naval Gunfire | | | Aviation | | | Engineering & Maintenance | | | Administration | | | Operations & Intelligence | | | Logistics | | | Services | | | Total | | |
|---|---|---|---|---|---|---|---|---|---|---|---|---|---|---|---|---|---|---|---|---|---|---|---|---|
| | Count (n) | | | | | | | | | | | | | | | | | | | | | | | |
| | M | F | Total | M | F | Total | M | F | Total | M | F | Total | M | F | Total | M | F | Total | M | F | Total | M | F | Total |
| Army | 1,458 | 2 | 1,460 | 554 | 18 | 572 | 834 | 55 | 889 | 600 | 58 | 658 | 692 | 45 | 737 | 680 | 74 | 754 | 1,320 | 266 | 1,586 | 6,138 | 518 | 6,656 |
| Navy | 711 | 62 | 773 | 667 | 9 | 676 | 499 | 6 | 505 | 230 | 46 | 276 | 277 | 22 | 299 | 294 | 15 | 309 | 654 | 139 | 793 | 3,332 | 299 | 3,631 |
| Air Force | 30 | ** | 30 | 1,082 | 24 | 1,106 | 672 | 42 | 714 | 373 | 67 | 440 | 719 | 99 | 818 | 390 | 55 | 445 | 731 | 232 | 963 | 3,997 | 519 | 4,516 |
| Marines | 208 | 2 | 210 | 229 | 4 | 233 | 128 | 0 | 128 | 240 | 3 | 243 | 98 | 4 | 102 | 214 | 2 | 216 | 42 | 1 | 43 | 1,159 | 16 | 1,175 |
| Total | 2407 | 66 | 2,473 | 2532 | 55 | 2,587 | 2133 | 103 | 2,236 | 1443 | 174 | 1,617 | 1786 | 170 | 1,956 | 1578 | 146 | 1,724 | 2747 | 638 | 3,385 | 14626 | 1352 | 15,978 |
| | Prevalence (per 1000 officers) | | | | | | | | | | | | | | | | | | | | | | | |
| Army | 7.60 | 1.13 | 7.54 | 6.40 | 3.78 | 6.26 | 6.75 | 2.70 | 6.17 | 11.22 | 2.92 | 8.97 | 9.80 | 3.33 | 8.76 | 9.73 | 3.69 | 8.38 | 8.48 | 4.10 | 7.19 | 8.16 | 3.57 | 7.42 |
| Navy | 8.13 | 6.20 | 7.93 | 6.85 | 1.52 | 6.55 | 8.91 | 1.87 | 8.53 | 11.34 | 8.33 | 10.69 | 9.73 | 4.81 | 9.05 | 13.31 | 4.29 | 12.08 | 8.02 | 3.30 | 6.41 | 8.48 | 4.00 | 7.76 |
| Air Force | 5.47 | ** | 5.47 | 6.83 | 2.91 | 6.63 | 8.85 | 3.54 | 8.14 | 11.21 | 5.07 | 9.46 | 9.54 | 5.76 | 8.84 | 10.04 | 5.41 | 9.08 | 7.45 | 4.13 | 6.24 | 8.23 | 4.44 | 7.50 |
| Marines | 5.28 | 4.69 | 5.27 | 5.41 | 3.23 | 5.35 | 5.83 | 0.00 | 5.55 | 12.03 | 1.02 | 10.62 | 5.23 | 3.18 | 5.10 | 8.71 | 0.66 | 7.83 | 8.38 | 1.45 | 7.54 | 6.74 | 1.50 | 6.43 |
| Total | 7.42 | 5.41 | 7.35 | 6.58 | 2.73 | 6.39 | 7.69 | 2.82 | 7.12 | 11.36 | 4.19 | 9.59 | 9.24 | 4.65 | 8.51 | 10.15 | 3.97 | 8.97 | 8.07 | 3.89 | 6.71 | 8.11 | 3.89 | 7.43 |
| Occupation Codes | O205: Ground and Naval Arms; O206: Missiles | | | O201: Fixed-Wing Fighter/Bomber Pilots; O202: Other Fixed-Wing Pilots; O203: Helicopter Pilots; O204: Aircraft Crews | | | OFF4: Engineering and Maintenance Officers | | | OFF1: General Officers and Executives, N.E.C.; OFF7: Administrators | | | O207: Operations Staff; O301: General Intelligence; O302: Communications Intelligence; O303: Counterintelligence | | | OFF8: Supply, Procurement and Allied Officers | | | OFF5: Scientists and Professionals; OFF6: Health Care Officers | | | All Officer Specialties | | |

Supplemental Table 2. Count and prevalence of Achilles tendon ruptures among enlisted members in the US Armed Forces, 2006-2018

| Enlisted Specialty | Special Operations Forces | | | Infantry | | | Mechanized/ Armor | | | Artillery/Gunnery | | | Aviation | | | Engineers | | | Maintainance | | | Administration, Intelligence, & Communication | | | Logistics | | | Maritime/Naval Specialties | | | Total | | |
|---|---|---|---|---|---|---|---|---|---|---|---|---|---|---|---|---|---|---|---|---|---|---|---|---|---|---|---|---|---|---|---|---|---|
| Count (n) | | | | | | | | | | | | | | | | | | | | | | | | | | | | | | | | | |
| | M | F | Total | M | F | Total | M | F | Total | M | F | Total | M | F | Total | M | F | Total | M | F | Total | M | F | Total | M | F | Total | M | F | Total | M | F | Total |
| Army | 262 | ** | 262 | 2,703 | ** | 2,703 | 366 | ** | 366 | 1,262 | 39 | 1,301 | ** | ** | ** | 484 | 3 | 487 | 5,080 | 352 | 5,432 | 7,125 | 1,166 | 8,291 | 3,356 | 539 | 3,895 | 53 | 6 | 59 | 20,691 | 2,105 | 22,796 |
| Navy | 47 | ** | 47 | ** | ** | ** | ** | ** | ** | 102 | 9 | 111 | 193 | 18 | 211 | ** | ** | ** | 6,127 | 535 | 6,662 | 3,315 | 457 | 3,772 | 1,119 | 148 | 1,267 | 612 | 92 | 704 | 11,515 | 1,259 | 12,774 |
| Air Force | ** | ** | ** | ** | ** | ** | ** | ** | ** | ** | ** | ** | 296 | 14 | 310 | ** | ** | ** | 7,245 | 341 | 7,586 | 5,247 | 1,088 | 6,335 | 2,145 | 275 | 2,420 | ** | ** | ** | 14,933 | 1,718 | 16,651 |
| Marines | ** | ** | ** | 761 | ** | 761 | 35 | ** | 35 | 83 | ** | 83 | 41 | 3 | 44 | 86 | 0 | 86 | 2,071 | 88 | 2,159 | 2,671 | 234 | 2,905 | 871 | 77 | 948 | ** | ** | ** | 6,619 | 402 | 7,021 |
| Total | 309 | ** | 309 | 3464 | ** | 3464 | 401 | ** | 401 | 1447 | 48 | 1495 | 530 | 35 | 565 | 570 | 3 | 573 | 20523 | 1316 | 21,839 | 18358 | 2945 | 21,303 | 7491 | 1039 | 8,530 | 665 | 98 | 763 | 53,758 | 5,484 | 59,242 |
| Prevalence (per 1000 enlisted members) | | | | | | | | | | | | | | | | | | | | | | | | | | | | | | | | | |
| Army | 4.25 | ** | 4.25 | 4.16 | ** | 4.16 | 4.46 | ** | 4.46 | 6.12 | 4.76 | 6.07 | ** | ** | ** | 4.30 | 2.09 | 4.27 | 5.88 | 4.34 | 5.75 | 8.02 | 4.66 | 7.29 | 6.94 | 5.20 | 6.64 | 12.27 | 11.59 | 12.19 | 6.17 | 4.73 | 6.00 |
| Navy | 3.55 | ** | 3.55 | ** | ** | ** | ** | ** | ** | 4.11 | 2.33 | 3.87 | 4.39 | 3.98 | 4.36 | ** | ** | ** | 5.34 | 3.48 | 5.12 | 8.36 | 3.80 | 7.30 | 7.24 | 3.98 | 6.61 | 6.22 | 4.05 | 5.81 | 6.13 | 3.68 | 5.75 |
| Air Force | ** | ** | ** | ** | ** | ** | ** | ** | ** | ** | ** | ** | 6.82 | 5.71 | 6.76 | ** | ** | ** | 7.97 | 5.30 | 7.80 | 10.75 | 4.94 | 8.94 | 8.54 | 5.18 | 7.95 | ** | ** | ** | 8.83 | 5.05 | 8.20 |
| Marines | ** | ** | ** | 2.50 | ** | 2.50 | 1.41 | ** | 1.41 | 2.39 | ** | 2.39 | 2.30 | 5.61 | 2.39 | 2.31 | 0.00 | 2.22 | 4.95 | 4.03 | 4.90 | 7.85 | 4.34 | 7.37 | 4.82 | 4.12 | 4.76 | ** | ** | ** | 4.87 | 4.17 | 4.83 |
| Total | 4.13 | ** | 4.13 | 3.63 | ** | 3.63 | 3.75 | ** | 3.75 | 5.45 | 3.98 | 5.38 | 5.04 | 4.66 | 5.01 | 3.80 | 1.00 | 3.75 | 6.15 | 4.10 | 5.97 | 8.69 | 4.57 | 7.73 | 7.00 | 4.88 | 6.65 | 6.47 | 4.22 | 6.06 | 6.49 | 4.48 | 6.23 |
| Occupation Codes | E011: Special Forces | | | E010: Infantry, General | | | Occupation: E020: Armor and Amphibious, General | | | E041: Artillery and Gunnery; E042: Rocket Artillery; E043: Missile Artillery, Operating Crew | | | E050: Air Crew, General; E051: Pilots and Navigators | | | E030: Combat Engineering, General | | | ENL7: Craftsworkers; ENL1: Electronic Equipment Repairers; ENL6: Electrical/ Mechanical Equipment Repairers | | | ENL2: Communications and Intelligence Specialist; ENL5: Functional Support and Administration | | | ENL8: Service and Supply Handlers | | | E062: Small Boat Operators; E063: Seamanship, General; E060: Boatswains | | | All Enlisted Specialities | | |

Supplemental Table 3. Comparison of the negative binomial regression models, with and without adjustment for zero inflation, assessing sex, age, rank, service branch, and year on the prevalence of Achilles tendon ruptures in the US Armed Forces, 2006-2015

| | Adjusted (Hurdle) Model | | | | | | Unadjusted (Standard) Model | | |
|---|---|---|---|---|---|---|---|---|---|
| | Zero Model | | | Count Model | | | | | |
| | | 95% CI | | | 95% CI | | | 95% CI | |
| | OR | LL | UL | PR | LL | UL | PR | LL | UL |
| **Intercept** | $6.01*10^9$ | 0.00 | ∞ | **4.57** | **3.61** | **5.80** | **6.15** | **4.71** | **8.06** |
| **Sex:** Female | **0.16** | **0.09** | **0.29** | **0.71** | **0.65** | **0.79** | **0.65** | **0.58** | **0.72** |
| **Age, y**: 20-29 | 0.00 | 0.00 | ∞ | 0.92 | 0.75 | 1.14 | **0.75** | **0.59** | **0.95** |
| 30-39 | 0.00 | 0.00 | ∞ | **1.81** | **1.47** | **2.23** | 1.22 | 0.97 | 1.54 |
| 40+ | 0.00 | 0.00 | ∞ | **2.50** | **2.01** | **3.11** | **1.47** | **1.15** | **1.87** |
| **Rank:** Senior enlisted | **47.76** | **16.20** | **140.79** | 0.97 | 0.85 | 1.12 | **1.30** | **1.11** | **1.52** |
| Junior officer | **5.19** | **2.62** | **10.28** | 0.92 | 0.79 | 1.06 | 1.14 | 0.97 | 1.33 |
| Senior officer | **8.13** | **3.77** | **17.53** | **0.76** | **0.65** | **0.89** | 0.95 | 0.79 | 1.13 |
| **Service branch:** US Navy | **0.16** | **0.06** | **0.42** | 1.05 | 0.92 | 1.19 | 0.94 | 0.81 | 1.09 |
| US Marine Corps | **0.02** | **0.01** | **0.06** | **1.38** | **1.20** | **1.58** | 0.98 | 0.84 | 1.14 |
| US Air Force | **0.24** | **0.08** | **0.66** | **1.17** | **1.03** | **1.33** | 1.07 | 0.92 | 1.24 |
| **Year:** 2007 | 0.84 | 0.26 | 2.69 | 0.92 | 0.75 | 1.13 | 0.93 | 0.74 | 1.17 |
| 2008 | 0.71 | 0.22 | 2.24 | 0.94 | 0.76 | 1.15 | 0.93 | 0.73 | 1.17 |
| 2009 | 0.47 | 0.15 | 1.43 | 0.91 | 0.74 | 1.12 | 0.85 | 0.67 | 1.08 |
| 2010 | 0.56 | 0.18 | 1.75 | 0.82 | 0.66 | 1.01 | **0.78** | **0.61** | **0.98** |
| 2011 | 1.43 | 0.40 | 5.12 | **0.79** | **0.64** | **0.97** | 0.82 | 0.64 | 1.03 |
| 2012 | 0.63 | 0.20 | 1.98 | 0.88 | 0.71 | 1.08 | 0.84 | 0.66 | 1.06 |
| 2013 | 0.65 | 0.20 | 2.08 | 0.94 | 0.77 | 1.16 | 0.92 | 0.73 | 1.17 |
| 2014 | 0.56 | 0.18 | 1.75 | 0.98 | 0.79 | 1.20 | 0.95 | 0.75 | 1.19 |
| 2015 | 1.43 | 0.40 | 5.12 | 1.08 | 0.88 | 1.32 | 1.08 | 0.86 | 1.36 |
| Negative log-likelihood ratio | | | 2358 | | | | | 5075 | |

Bolded values indicate statistical significance ($p < 0.05$); CI, confidence interval; LL, lower limit; OR, odds ratio; PR, prevalence ratio; UL, upper limit. The zero model employed a binomial regression to assess groups with positive counts versus no outcomes. In the second step, the count model employed a truncated negative binomial regression (omitting zero counts) based on the log link. Reference groups: sex (male), age (18-19), rank (junior enlisted), service branch (US Army), year (2006).
